# Performance Assessment of First-Generation Anti-SARS-CoV-2 Serological Assays

**DOI:** 10.1101/2020.09.22.20197046

**Authors:** Mehjabeen Imam, Shabnum Khawaja, Arshi Naz, Ahson Siddiqui, Tehmina S. Nafees, Amber Younus, Usama Shamsi, Imran Shabir, Shakir Ahmed, Naveen Tariq, Salman Tariq, Tahir S. Shamsi

**Author notes:** **Corresponding Author** Tahir S. Shamsi, MBBS, FRCP(London), FRCP (Edin), MRCPath (UK), FRCPath (London), Dean, Postgraduate Studies, Professor and Chairman, Department of Internal Medicine, Consultant Haematologist & Transplant Physician. Professor & Convenor. Children Hospital Lahore & National Institute of Blood Diseases & Bone Marrow Transplantation Karachi Country Advisor, Royal College of Pathologists, UK.

## Abstract

The clinical and epidemiological use of SARS-CoV-2 antibody assays is under debate with urgent need to validate and verify the performance of SARS-CoV-2 serologic assays. We aim to assess the clinical and analytical performance of three commercial serological assays of SARS-CoV-2, comparing three anti-SARS-CoV-2-IgG ELISA and identifying the seroconversion and seroprevalence in our population.

A cross sectional study conducted from April 2020 to July 2020 at National Institute of Blood disease and Bone Marrow Transplantation Karachi, Pakistan with sample size of 404, enrolled consecutively. Participants were categorized into four groups’ namely convalescent plasmadonors (CPDs n=239), health care professionals (HCPs n=44), healthy blood donors (HBDs n=70) and from community (n=51).

We evaluated the performance of Elecsys anti-SARS-CoV-2 electrochemiluminescence (ECLIA) assay on Cobas-e411 by Roche, three qualitative anti-SARS-CoV-2-IgG enzyme linked imunosorbant assay (ELISA) by (Generic assays, Euroimmun & Omega diagnostics), one quantitative ELISA assay by AESKU Diagnostics and two immune chromatography(ICT) kits namely InstaTest™ by CORTEZ and TEST IT by TURKLAB.

From total 404 subjects, 322 (83.5%) were males. Mean age was 36.79±11.95 years. Among 239 in CPDs group, 202(84.5%) showed positive antibodies by ECLIA. The qualitative anti-SARS-CoV-2 IgG ELISA was positive in 174 (72.8%) and quantitative IgG in 180(75.3%) with mean titer of 56.7 ±39.7 U/ml. Sensitivity and specificity of ECLIA were 97.44& 99%, ELISA by Generic assays were 67.85% and 89.9%; Euroimmun had 90.38% and 94.9%; Omega Diagnostics 96.4% and 95% and the AESKULISA 93.75% and 100% respectively. Seroconversion was found to be 53.8% and 77.77% within 7 -8 days and 12 to 14 days post onset of symptoms respectively. ICT had more specificity but less sensitivity. Seroprevalence was found to be 84.5%, 40.9% and 21.4% in CPDs, HCPs and HBDs respectively.

The Roche ECLIA, qualitative ELISA by Omega Diagnostics & Euroimmun showed higher sensitivity as well as higher specificity. Quantitative ELISA has higher specificity and relatively high sensitivity. Significant numbers of COVID patients do not have detectable antibodies by all assays.

## Introduction

The global pandemic of COVID-19 caused by Severe Acute Respiratory Syndrome Corona virus 2 (SARS-CoV-2) affected Pakistan in mid-March^1^ and till mid-July 2020, a total of 254,000 cases were diagnosed and 5,500 (2.1%) deaths reported^2^. According to the Center of Disease Control (CDC), the recommended method for the diagnosis of COVID-19 is to detect the RNA of SARS-CoV-2 by real time Reverse Transcriptase Polymerase Chain Reaction (RT-PCR) with samples collected from nasopharynx and / or oropharynx^3, 4^.Commercial manufacturers developed serological testing kits using different methodologies^5^.FDA gave approval for the use of these kits as Emergency Use Authorization (EUA)^6^. Verification and validation of these assays is required to achieve the clinical accuracy of test results^7, 8^.

There are several advantages of serological testing. Pre analytical and analytical parts of serological assays have comparatively easy sample collection, simple to analyzeand requires less technical expertise as compared to nucleic acid detection. Serological testing can be performed in basic and small clinical laboratory settings^8,9^. Post analytical important uses of antibody testing include; to determine the exposure of this disease among tested population, to assess the immune response against this virus and to identify the actual number of people who had acquired this virus but were asymptomatic therefore they didn’t get tested on RT-PCR. The results of this testing will be significant for epidemiological studies, disease surveillance and later in monitoring of response to the vaccine^9, 10^. It plays vital role for identification of convalescent plasma donors for therapeutic use in treating moderate to severely ill COVID-19 patients^10^. There are several methodologies available for the antibody testing which include immune chromatographytechnique(ICT),electrochemiluminescenceimmunoassay(ECLIA),chemiluminee (CLIA) and enzyme linked immunosorbant assay (ELISA)^8,11^ etc. To-date very little data available about clinical and analytical performance of SARS-CoV-2 antibody assays; we planned to analyze the clinical and analytical performance differentCommercial Serological assays of SARS-CoV-2, compared three commercial anti-SARS-CoV-2-IgG ELISA assays and to see the Seroconversion of anti-SARS-CoV-2 in four different population groups.

## MATERIAL &METHOD

### Study Design

A cross sectional study conducted from April 2020 to June 2020 in Karachi, Pakistan after the approval from institution’s ethical review committee. Informed written consent was taken from all study subjects.Non-probability consecutive sampling technique was used for sampling. Three milliliter (ml) blood was collected in gel vacutainer from all subjects or serological testing and nasopharyngeal swab was taken for anti-SARS-CoV-2 RT-PCR testing. We divided our subjectsin following four groups shown in Table no 1. The anti-SARS-CoV-2 antibodies were tested by ECLIA, ICT and by ELISA. The ECLIA and ICT were qualitative while ELISA was performed on both qualitative as well as quantitative kits.

**Table No. 1:** Inclusion Criteria of our study.

| **<u>No</u>** | **<u>Group</u>** | **<u>Inclusion criteria</u>** |
| --- | --- | --- |
| 01 | Convalescent plasma donors (CPDs) for COVID-19 | Adult corona survivors of either gender aged 18 to 60 years, with no comorbidities, fully recovered from COVID-19 for at least two weeks<sup>12</sup>. |
| 02 | Health care professionals (HCPs) | Hospital staff of either gender aged 18 to 60 years who were experiencing symptoms associated with COVID-19 including fever, dry cough, body aches, flu-like symptoms, sore throat, new loss of taste or smell, diarrhea and difficulty in breathing<sup>13</sup>. Their PCR was done along with serological testing. |
| 03 | People from | Convalescent plasma donors spread the awareness and urged their |
|  | community | closed acquaintances to get them tested for COVID-19, along with them walk-in patients who were tested for PCR and anti SARS-CoV-2 IgG simultaneously were also taken in this group. |
| 04 | Healthy blood donors (HBDs) | Regular blood donors were recruited in this group and after their consent we tested them for anti-SARS-CoV-2 antibodies. |

### Analytical Performance Methodolog

For analytical performance, repeatabilitydone using known negative samples, pre-pandemic samples collected from blood bank archive, and known positive patient samples. Intra-assay precision was assessed by 20 times single run. Inter-assay precision was assessed by running these samples 20 times on separate runs on each method. Sensitivity of ECLIA was calculated on COVID-19 RT-PCR positive patients tested after 2-5 days of onset of symptoms. They were closely followed up; 13 individuals were tested for antibody presence on 7 - 8 days post onset of symptoms and 18 study subjects were checked after 12-14 days after symptoms. Sensitivity at more than 28 days post symptoms was checked in CPDs’ samples. The sensitivity of qualitative and quantitative ELISA and ICTwas checked on CPDs’samples.The number of participants in each group is shown in Table no 2.

**Table No. 2:** Groups of samples in study.

| S. No. | Name of group | Number of samples |
| --- | --- | --- |
| 1 | Convalescent plasma donors | 239/404 |
| 2 | Community | 51/404 |
| 3 | Health care professionals | 44/404 |
| 4 | Healthy blood donors | 70/404 |

### Electro-Chemiluminescence immunoassay (ECLIA)

Total antibody against SARS-CoV-2(including IgG, IgM and IgA) detected by using double-antigen sandwich assay on Cobas 1Immunoassay analyzer(Roche diagnostics International Ltd atRotkreuz Switzerland).The assay used a recombinant protein representing the nucleocapsid (N) antigen. Result reported as Reactive = if Cut of Index (COI)>1.0 and Non-Reactive =COI<1.0.

### Immune Chromatography Technique (ICT Method)

The anti-SARS-CoV-2 diagnosis qualitative detection of IgG and/or IgM based on the principle of lateral flow chromatography. Two kits were used for this ICT method namely, Insta Test™ 2019-nCoV IgG/IgM Rapi Card Insta Test™, manufactured by CORTEZ Diagnostics, Inc. Woodland Hills California USA and TEST IT SARS-COV-2 IGM/IGG AB TEST by TURKLAB Medical Devices, Menderes Izmir Turkey.

### Enzyme linked immunosorbent assay (ELISA)

This technique is used to detect anti-SARS-CoV-2IgG antibody in subject’s serum. There were three kits for qualitative ELISA namely, Generic Assays GA CoV-2 IgG by GA Generic Assays GmbH,Dehlewitz, Germany, anti SARS-CoV-2 ELISA (IgG) by EUROIMMUN Medizinische Labordiagnostika AG, Lubeck Germany and Omega Diagnostics Covid-19 IgG ELISA Kit by Generic Diagnostics Ltd, Cambridgeshire, UK. The only quantitative kit of anti-SARS-CoV-2IgG available till date is AESKULISA® SARS-CoV-2 NP IgGmadeby AESKU. DIAGNOSTICS GmbH & Co. KG, Wendelsheim Germany.All of these kits detected antibodies against Nucleocapsid protein.

### STATISTICAL ANALYSIS

Mean and Standard deviation for quantitative variables, while frequency with percentage for categorical variable was calculated. In repeatability mean, SD and a % coefficient of variation (%CV) was calculated. For electrochemiluminescenceassay %CV calculated by using numerical COI. For qualitative anti-SARS-CoV-2 IgG ELISA, the %CV was calculated by using numerical optical density (OD), while for quantitativeIgG-ELISA%CV was calculated by using OD and U/mL. The sensitivity, specificity, positive predictive valueand negative predictive value for each assay was calculated.Analysis was done on statistical package for social science SPSS (Version 23).

## RESULTS

Total 404 subjects were recruited in the study. Out of these, 342 (84.6%) were males.Mean age of the subjects was 36.79±11.95 years. There were 239 CPDs; of them, 202 (84.5%) had a positive anti-SARS-CoV-2 by ECLIA.The qualitative ELISA performed in 174/239 CP donors, found to be positive in 142/174(82%) with a mean serum dilution of >1:160 (Range >1:80 to >1:320 serum dilution).Quantitative IgG by ELISA performed on total 252 subjects, of these 239(94.8%) were CPDs; 180 of these 239 (75.3%) were positive for IgG and their mean anti-SAR-CoV-2IgG level was 56.7±39.7U/ml. Seroprevalence was found to be 84.5% in CPDs at 21 days post onset of symptoms.

A total of 44 HCPs were recruited, their RT-PCR performed 3 to 5 days after the onset of symptoms. Their anti-SARS-CoV-2 antibody levels were tested by ECLIAon 7-8days and 12-14 days;seven of 13(53.8%) developed anti-SAR-CoV-2 within 10 days while 14 of the 18 (77.77%) developed antibodies by the end of second week of symptoms. Seroprevalence was found to be 40.9% in HCPs at 21 days post onset of symptoms.

Of the 70 HBDs screened, 15(21.4%) developed this antibody. These 15were also analyzed by quantitative IgGELISA and 14(93.3%) of them showed an antibody level of 27.2 ±19.95 U/ml. Seroprevalence was found to be 21.4% in asymptomatic HBDs.

In community group 12 patients were PCR positive and out of these 12, three patients had antibodies positive on ECLIA. Seroprevalence could not be calculated in this group since these patients were lost to follow up.

Analytical Performance of Serological Assaysshown in table no. 3.

**Table No. 3:** Intra- and inter-assay Repeatability of ECLIA& ELISA.

| **ASSAYS** | **Positive Control (%CV)** | **Negative Control (%CV)** |
| --- | --- | --- |
| **ECLIA (COI)** | 3.05±0.24 (7.86%) | 0.09±0.002 (2.53%) |
| **Generic Assays (OD)** | 2.46 ± 0.10 (4.0%) | 0.099 ±0.004 (3.6%) |
| **Euroimmun (OD)** | 1.34 ± 0.06 (4.50%) | 0.020 ±0.001 (5.00%) |
| **Omega Diagnostics (OD)** | 1.01±0.05 (4.8%) | 0.369 ±0.017 (4.35%) |
| **AESKULISA (OD)** | 1.69±0.03 (1.7%) | 0.12 ±0.01 (5.4%) |
| **AESKULISA (U/mL)** | 58.84±2.87 (5.01%) | 3.23 ±0.24 (2.1%) |

The diagnostic performance(sensitivity, specificity, PPV & NPV) of all the serological assays shown in table no. 4.

**Table No. 4:**
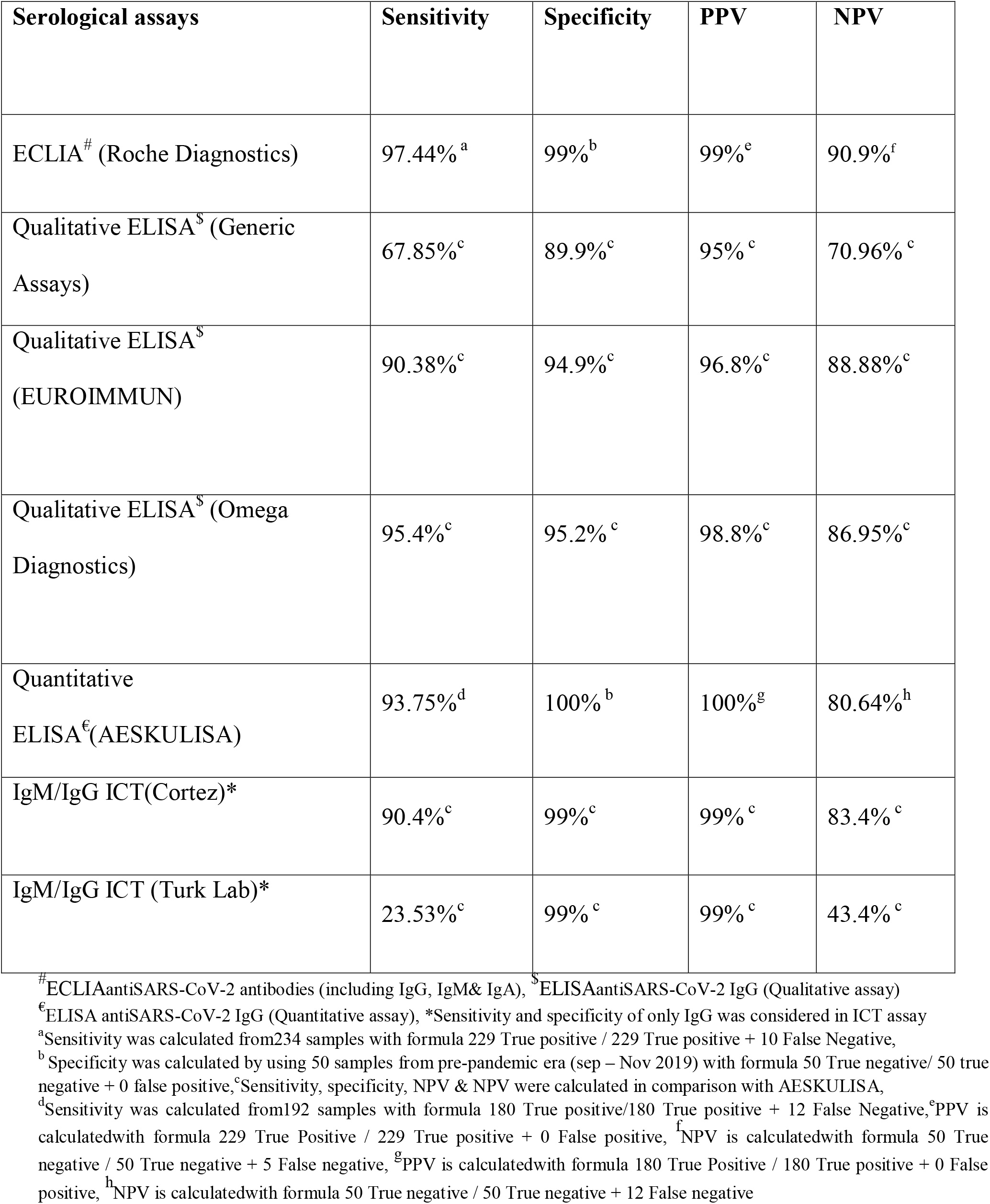
Diagnostic performance of all the serological assays used in the study.

## DISCUSSON

We evaluated the performance of three different serological assays infourdifferentgroups. This study included convalescent plasma donors’whorecovered from COVID-19 infection.The performance characteristics of different kits e.g., sensitivity claimed by their manufacturers, fell short since our calculated sensitivities were below thatofthe manufacturers’ claim. This finding is in accordance with an Australian report published on 29^th^ April 2020 that assessed post-market validation of three serological assays for COVID-19 where they tested the serological assays on CPDs and found out their own diagnostic performance for each kit^14^.

Out of the three methodologies, ECLIA assay had sensitivity 97.44%, qualitative ELISA by Omega diagnostics had 96.4% while other two qualitative ELISA had lower sensitivity. This may be due to the fact that electrochemiluminescence assay detects total antibodies (including IgG) against SARS-CoV-2 while ELISA is only IgG specific.

Mei San Tang et al compared Abbott Chemiluminescenceassay and ELISA within 5 days of onset of symptoms, none of the immunoassays was able to detect the antibodies^9^. This leads to an important observation that when to test for the presence of antibodies against SARS-CoV-2 antibodies in our population.The samples which were taken from HCPs and community within 3 to 5 days post the onset of symptoms did not show antibody positivity on either assay. This finding is in contrast with the manufacturer’s claim that for ECLIA by Roche which showed 65.5% (CI 56%-74%) sensitivity when tested within 6 days PCR confirmation. None the less, gold standard for the diagnosis of acute SARS-CoV-2 infection is RT-PCR and we will need larger cohorts of patients to identify correct time for using serological assays in our population.

We found important and interesting finding in our study, out of 239 CPDs the 37 (15.5%) did not develop antibodies against SARS-CoV-2 virusbyany of the testing method i.e. ECLIA, quantitative and qualitative ELISA.Cellular immunity might have been developed in these seronegative CP donors. This remains to be tested in our cohort. Shane et al in June2020 detected CD4+ and CD8+ T cells in 100% and 70% of convalescent COVID patients; they further discovered that T cell responses are not only on spike (S) but also on M, N, and other ORFs of SARS-CoV-2^15^.

The two rapid testing devices using ICT methodology showed the sensitivity for IgM/IgG tests ranging from 23.53% to 90.4%. The specificity of both the devices on IgM and IgG tests was found to be 95% to 100%. The results of sensitivity are, however in contrast with the findings of Z.Zainol Rashid et al. They used nine rapid devices based on the principle of colloidal gold-labeled immunochromatography. Their sensitivity for both IgM and IgG tests ranges between 72.7% and 100%, while specificity ranges between 98.7% to 100%^16^.

The heathyblood donors had no symptoms of COVID-19 infection, they were healthy and active;a quarter of them seroconverted against COVID-19. This is a significant finding as it highlights the prevalence of this disease in general population. This observation has not been reported until now. We have also correlated the results of ECLIA and ELISA in our groups of CP donors and healthy blood donors which is showing positive correlation between these two assays.

Limitation of study was that we could not perform RT-PCR of healthy blood donors and ELISA of health care professionals and people from community due to cost limitation.

The strength of the study was that weincluded 239 convalescent plasma donors who recovered from COVID-19 infection that is by far the largest number of convalescent plasma donors recruited in any study.Wedid quantitative ELISA in the CPDs’ sample; this is the largest number of CPDs tested on quantitative ELISA till today in Pakistan.

## CONCLUSION

In conclusion, the Roche ECLIA assay has higher sensitivity then ELISA and ICT methods in our study. ECLIA and ELISA are not to be used for the acute diagnosis of SARS-CoV-2 infection since these assays have poor sensitivity when tested within 7 days post onset of symptoms. Serological assays are important in determining the prevalence of this disease. In our study, all the commercially available serological assays are detecting antibodies against nucleocapsid protein of the SARS-CoV-2 virus. New serological assays should also be evaluated for the detection of antibodies against receptor binding domain of spike protein that is said to be neutralizing antibody^17^. Serial monitoring of IgG titers among patients for the collection of CP as well as pooled human IVIG is also an important query for the future of treatment of COVID-19. Measuring immunity to SARS-CoV-2 is essential for vaccinedevelopment, T cell responses are not only on spike but also on M, N, and other ORFs. These newer methods are being available for measure the T-Cell response, need of validate these assays is highly recommended.

## Data Availability

There is no external material online or any external datasets are used in this manuscript. This is a single center study. Manuscript has original content.

## ETHICS APPROVAL AND CONSENT TO PARTICIPATE

This study was starts after the approval of Institutional Review Board (IRB)/Ethical Committee of Hospital.

## AVAILABILITY OF DATA AND MATERIALS

Data is confidential.

## FUNDING OR FELLOWSHIP GRANT

None

## ACKNOWLEDGEMENTS

All authors thankful to participants, on the basis of their results we will be able to choose best method for the detection of SARS-CoV-2antibodies in future.

## LIST OF ABBREVIATIONS

SARS-CoV-2: Severe Acute Respiratory Syndrome Coronavirus-2
COVID-19: Coronavirus disease of 2019
IgG: Immunoglobulin G
ELISA: Enzyme Linked Immunosorbant Assay
CPDs: Convalescent Plasma Donors
HCPs: Health Care Professionals
HBDs: Healthy Blood Donors
ECLIA: Electro ChemiluminescenceImmunoassay
ICT: Immune Chromatographic Techinique
CDC: Centre of Disease Control and Prevention
RT-PCR: Reverse Transcriptase-Polymerase Chain Reaction
FDA: Food and Drug Administration
EUA: Emergency Use Authorization
CLIA: ChemiluminescenceImmunoassay
IgM: Immunoglobulin M
IgA: Immunoglobulin A
COI: Cut Off Index
OD: Optical Density
U/mL: Units per milliliter
SPSS: Statistical Package for the Social Sciences
PPV: Positive Predictive Value
NPV: Negative Predictive Value
CI: Confidence Interval
ORF: Open Reading Frame

